# Stability of diagnostic coding of psychiatric outpatient visits across the transition from the second to the third version of the Danish National Patient Registry

**DOI:** 10.1101/2022.03.03.22271695

**Authors:** Martin Bernstorff, Lasse Hansen, Erik Perfalk, Andreas Aalkjær Danielsen, Søren Dinesen Østergaard

## Abstract

**Background:** In Denmark, data on hospital contacts are reported to the Danish National Patient Registry (DNPR). The ICD-10 main diagnoses from the DNPR are often used as proxies for mental disorders in psychiatric research. With the transition from the second version of the DNPR (DNPR2) to the third (DNPR3) in February-March 2019, the way main diagnoses are coded in relation to outpatient treatment changed substantially. Specifically, in the DNPR2, each outpatient treatment course was labelled with only one main diagnosis. In the DNPR3, however, each visit during an outpatient treatment course is labelled with a main diagnosis. We assessed whether this change led to a break in the diagnostic time-series represented by the DNPR, which would pose a threat to the research relying on this source.

**Methods:** All main diagnoses from outpatients attending the Psychiatric Services of the Central Denmark Region from 2013 to 2021 (n=100,501 unique patients) were included in the analyses. The stability of the DNPR diagnostic time-series at the ICD-10 subchapter level was examined by comparing means across the transition from the DNPR2 to the DNPR3.

**Results:** While the proportion of psychiatric outpatients with diagnoses from some ICD-10 subchapters changed statistically significantly from the DNPR2 to the DNPR3, the changes were small in absolute terms (e.g., +0.9% for F2 - psychotic disorders and +0.1% for F3 - mood disorders).

**Conclusion:** The change from the DNPR2 to the DNPR3 is unlikely to pose a substantial threat to the validity of most psychiatric research at the diagnostic subchapter level.

**Data availability:** Due to restrictions in Danish law for protecting patient privacy, the data used in this study is only available for research projects conducted by employees in the Central Denmark Region following approval from the Legal Office under the Central Denmark Region (in accordance with the Danish Health Care Act §46, Section 2). However, similar data can be accessed through Statistics Denmark. Danish institutions can apply for authorization to work with data within Statistics Denmark, and such organisations can provide access to affiliated researchers inside and outside of Denmark.

**Significant Outcomes:**

- A significant administrative change in diagnostic coding of psychiatric outpatient treatment in Denmark did not induce marked destabilisation in the incidence of psychiatric diagnoses or the number of diagnoses received by each patient.
- In the DNPR3, most outpatient treatment courses are labelled with the same main diagnosis (subchapter level) at the first and last visit.

**Limitations:**

- Analyses were performed on data from the Central Denmark Region. While the administrative change was at the national level, and results should generalise, this is not testable from these data.
- Data was obtained from the Business Intelligence Office. While this source receives data from the same source as the National Patient Registry, and should be identical, this could not be tested.

## Introduction

Danish registries are widely used in psychiatric research.^1–3^ One such registry, the Danish National Patient Registry (DNPR), contains information on emergency treatment, admissions and outpatient visits for all Danish residents and all public hospitals in Denmark.^1^ In the DNPR, the registered main diagnosis—coded according to the International Classification of Diseases, 10th revision^4^—represents the condition leading to a specific hospital contact. The validity of the main diagnoses is well established and they are widely used as either exposure-, confounder- or outcome variables in psychiatric research.^1,5–7^

With the transition from the second version of the DNPR (DNPR2) to the third (DNPR3), which occurred in the period from February 2nd to March 3rd 2019 across the five Danish Regions, the way main diagnoses are coded in relation to outpatient treatment changed substantially (see Figure 1 for an illustration).^8^ Specifically, under the DNPR2 paradigm, all visits in a treatment course “inherited” the final main diagnosis, which should be representative of the entire treatment course. With the introduction of the DNPR3, however, coding instructions were updated. Since then, every single outpatient visit has been coded with a main diagnosis to cover that specific visit only. As a result, the main diagnosis under the DNPR3 is based on the—sometimes limited—clinical information available at the time of each visit, rather than the complete information available at the end of the treatment course.

**Figure 1.**
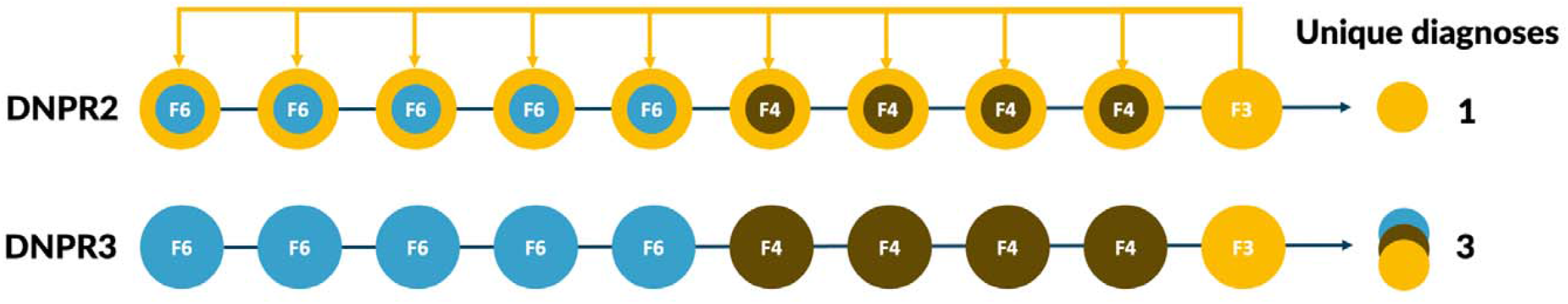
Number of unique diagnoses assigned for a psychiatric outpatient treatment course under the DNPR2 and DNPR3 eras, respectively During the DNPR2 era, when a series of visits was reported to the DNPR2, the final main diagnosis was used to overwrite the main diagnoses of all previous visits in the same treatment course. This was no longer the after the transition to the DNPR3, which can result in differences in the number of main diagnoses assigned for identical treatment courses. In this example, it results in more unique diagnoses pr. treatment course. In the example, F3, F4 and F6 refers to diagnostic categories in the mental disorder chapter of the ICD-10. F3: Mood disorders. F4: Neurotic, stress-related and somatoform disorders. F6: Disorders of adult personality and behaviour.

The change in diagnostic coding practice outlined above may have led to a destabilization/break in the diagnostic time-series represented by the DNPR, which would pose a significant threat to the many psychiatric research activities that rely on the main diagnoses, including our ongoing work based on data from the PSYchiatric Clinical Outcome Prediction (PSYCOP) cohort.^9^ Indeed, if patients are assigned a broader range of diagnoses in the DNPR3 compared to the DNPR2, the population-adjusted incidence of most diagnoses will go up, without reflecting a true increase in morbidity. However, whether such a destabilization has indeed occurred remains unexplored. Therefore, we examined the stability of diagnostic coding of psychiatric outpatient treatment across the DNPR2-DNPR3 transition at three levels: i) at the level of the psychiatric ICD-10 subchapters (e.g. F2 - psychotic disorders and F3 - mood disorders), ii) at the level of a connected series of outpatient visits, i.e., whether the diagnosis changed from the first to the last visit in an outpatient treatment course, and iii) at the level of the individual, i.e. whether the number of different diagnoses assigned to each patient was stable. Furthermore, we examined two strategies for maintaining comparability across the DNPR2-DNPR3 transition: Specifically, we recoded all visits in relation to an outpatient treatment course in the DNPR3 era with either the most *“severe”* diagnosis in the series of visits or with the *final* diagnosis in the series of visits, respectively.

## Methods

### Population and data

For the period from January 1, 2013, to June 1, 2021, we extracted the main diagnoses for all in- and outpatient visits to the Psychiatric Services of the Central Denmark Region – one of five Danish Regions – with a catchment population of approximately 1.3 million people covered by five psychiatric hospitals. Data was acquired from the Business Intelligence Office in the Central Denmark Region, resulting in the most up-to-date information available. The Business Intelligence Office and the DNPR receive data from the same source, namely the Electronic Health Record system used by all hospitals in the Central Denmark Region (MidtEPJ). This source contains data from the 30^th^ of May 2011 and onwards. However, as the psychiatric hospitals in the Central Denmark Region were gradually onboarded to MidtEPJ, the data from the first two years had unstable proportions of diagnostic codes that are unlikely to reflect general instability in the coding (see Supplementary Figure 1). Therefore, data from the time prior to January 1st 2013 were excluded for further analyses.

### Analyses

We only examined outpatient visits for the primary analyses, as the change in diagnostic coding should only affect the practice concerning this type of visits. We considered outpatient visits to be part of a connected treatment course in the DNPR3 era if they had the same unique course-element-identifier (in Danish DNPR3-terminology: “forløbselement-id”). The course-element-identifier covers patient-activities related to the same treatment course at the same department (for more details, see Supplementary Table 1). Furthermore, since the treatment courses defined by the course-element-identifier in the DNPR3 era are not directly comparable to the treatment courses as defined in the DNPR2 (see Supplementary Table 1), we also examined whether we could construct treatment courses for the DNPR3 era that more closely resemble those from the DNPR2. Specifically, we performed supplementary analyses in which all visits by the same patient to the same outpatient clinic in the DNPR3 era were considered part of the same treatment course.

We also examined two mitigation strategies regarding the stability of main diagnoses from each ICD-10 F-Subchapter (FX) across the transition from the DNPR2 to the DNPR3 eras. First, we coded all visits in a DNPR3 treatment course by the most “severe” diagnosis, defined as the FX diagnosis with the lowest digit, except F1 (Substance abuse), which was considered the least severe. Second, we coded all visits in a DNPR3 treatment course with the final diagnosis from the course, mimicking the DNPR2 approach.

#### i) Stability of main diagnoses from each ICD-10 F-Subchapter across the DNPR2-DNPR3 eras

As the main level of information in ICD-10 chapter F (mental and behavioural disorders) is captured by the first digit (e.g. F0: Organic mental disorders; F1: Substance use disorders; F2: Psychotic disorders), we used the main diagnosis at this level for the primary analyses. Visits with a main diagnosis of F99 (Mental disorder, not otherwise specified) were excluded, as they were rare (0.11% of all outpatient visits) and not conceptually related to the other F9-diagnoses (Behavioral and emotional disorders with onset usually occurring in childhood and adolescence). Visits were binned by 3-month periods from the transition date and rounded to the nearest beginning of a quarter (e.g. the 2nd of February 2019 was rounded to the 2nd of November 2018, whereas the 3rd of February 2019 was rounded to the 3rd of May 2019).

This ensured that the transition between the DNPR2 and the DNPR3 (2^nd^/3^rd^ of February 2019) was placed at the transition point between two bins.

Population-adjusted within-quarter incidences were calculated for each F-subchapter (FX) and each quarter using the equation below.

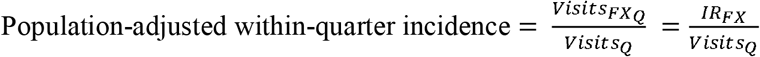

The numerator represents the number of patients with an outpatient visit that had a main diagnosis from the specific ICD-10 F-subchapter (FX), while the denominator represents the number of patients with any psychiatric outpatient visit in the same period. 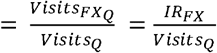 is the number of visits with a main diagnosis of FX during the quarter Q, *Visits*_*Q*_ is the number ofvisits during the quarter Q, and *IR*_*FX*_ is the incidence rate of FX for the quarter Q. As the number of patients are counted within a quarter, time is implicit.

For each FX, we calculated p-values for the difference between the mean proportion of visits belonging to the category before and after the transition from the DNPR2 to the DNPR3 using a t-test.

To ensure that the change in diagnostic coding practice did not spill over to admissions (inpatient hospital stays), we examined these as well, using the same approach as outlined above. Since the diagnostic coding of admissions should be less affected (if at all) by the transition from the DNPR2 to the DNPR3, as admissions were/are to be coded with one single main diagnosis both in the DNPR2 and DNPR3, we did not expect to see any breaks in the diagnostic time series for admissions.

#### ii) Diagnostic stability during outpatient treatment courses in the DNPR3 era

Based on the approach described above, the outpatient visits were grouped by their character-digit (FX) combination. Visits were considered part of a treatment course if they had the same unique course-element-identifier. To examine to which extent the first diagnosis in a DNPR3 sequence matched the last, we generated tables showing each first-/last-diagnosis combination at the FX level.

#### iii) Mean number of unique diagnoses per active outpatient treatment course across the DNPR2-DNPR3 eras

When counting main diagnoses at the level of the individual, diagnoses were truncated to the FXX-level. We ran sensitivity analyses with truncation at one (FX), three (FXX.X) and four (FXX.XX) digits. We also tested the difference between means before/after the DNPR2-DNPR3 transition using a t-test. Here, we considered a treatment course to be terminated 180 days after the last registered visit.

The threshold for statistical significance was set at .05. All p-values were corrected for multiple comparisons using the method of Benjamini and Hochberg.^10^ All analyses were carried out using R (www.r-project.org) and the code used for the analyses is available at: https://github.com/Aarhus-Psychiatry-Research/diagnostic-stability-lpr2-lpr3/

### Ethics

This study was carried out to ensure the validity/stability of the data used for studies based on the PSYCOP cohort.9 The use of electronic health records from the Central Denmark Region was approved by the Central Denmark Region Legal Office per the Danish Health Care Act §46, Section 2. According to the Danish Committee Act, ethical review board approval is not required for studies based solely on data from electronic health records (waiver for this project: 1-10-72-1-22). All data were processed and stored in accordance with the European Union General Data Protection Regulation and the project is registered on the internal list of research projects having the Central Denmark Region as the data steward.

## Results

Data from 100,501 outpatients with a total of 2,331,427 outpatient visits were analysed. The median age of the patients at the time of the first contact to the psychiatric services in the period from January 1st 2013 to June 1st 2021 was 27.6 years (25-75 percentiles: 17.0; 47.0 years) and 51.4% of the patients were women. The median number of outpatient visits per patient was 11 (25-75 percentiles: 4; 25).

### i) Stability of main diagnoses from each ICD-10 F-Subchapter across the DNPR2-DNPR3 eras

Table 1 and Figure 2 show that while the transition from the DNPR2 to the DNPR3 was associated with statistically significant changes in diagnostic proportions for most ICD-10 subchapters (FX), for all subchapters, these changes were small/modest on an absolute scale (e.g., +0.9% for F2 - psychotic disorders and +0.1% for F3 - mood disorders, respectively). Most diagnostic subchapters had trends in their frequency prior to the transition, and it appears from Figure 2B that only F8 (developmental disorders incl. autism) experienced a break in the time-series, which was, however, small in absolute terms (Figure 2A).

**Figure 2.**
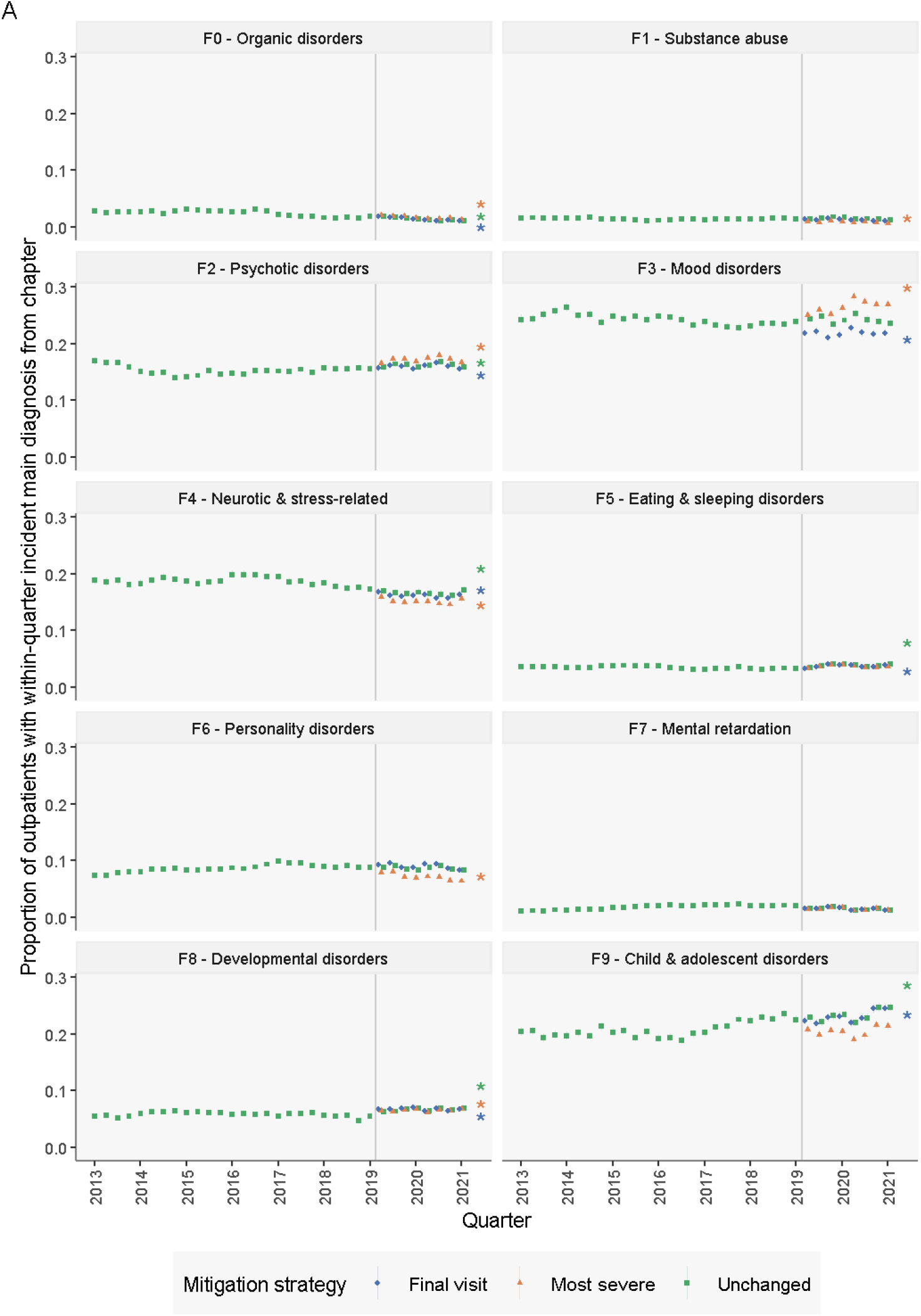
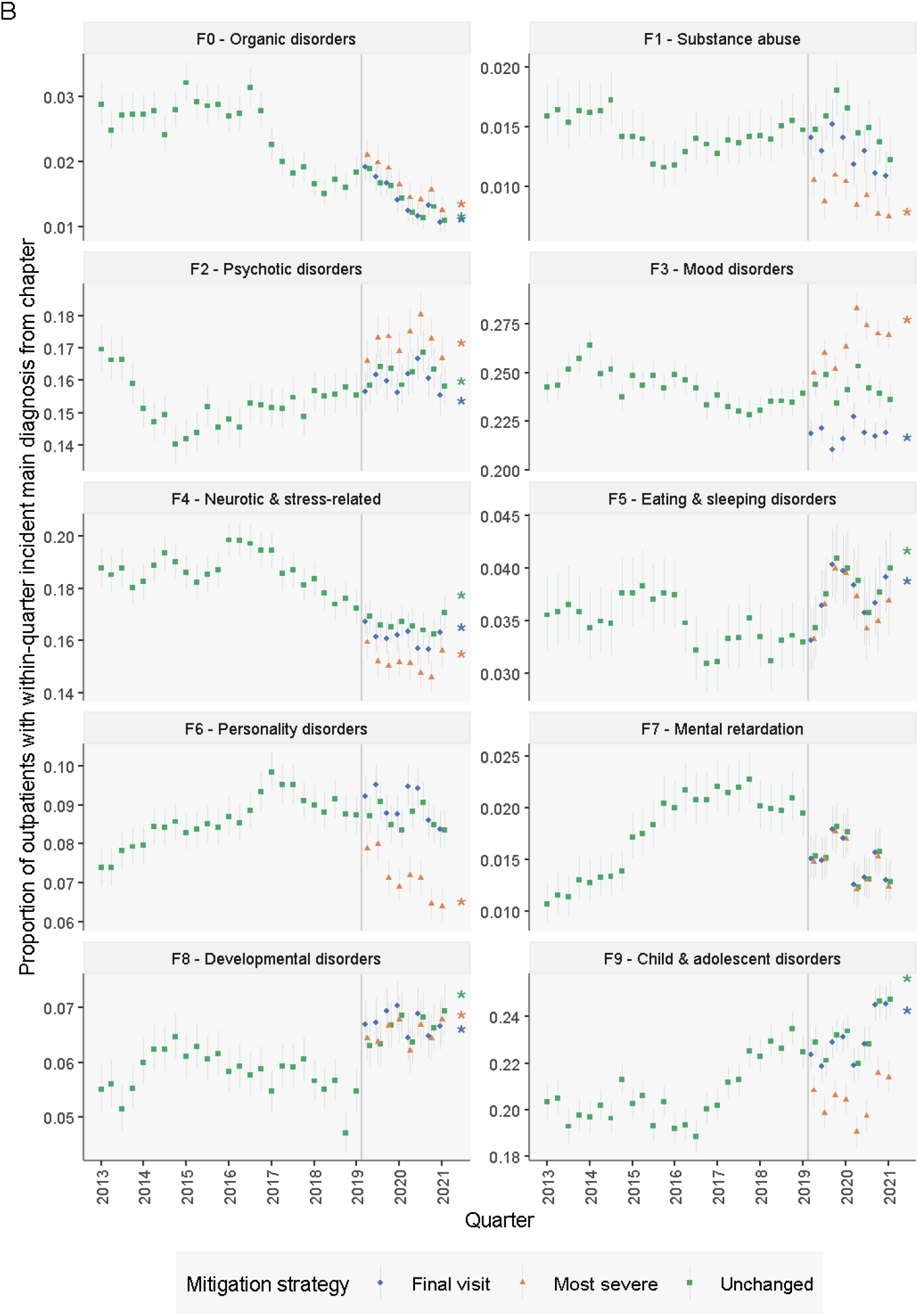
Proportion of outpatients in each quarter with a within-quarter incident main diagnosis by ICD-10 subchapter. A: y-scale is standardised across panels. B: y-scale is allowed to vary between panels. Proportion of all outpatients in each quarter that received a main diagnosis from each ICD-10 F-subchapter. Line ranges reflect 95% confidence intervals. The date of transitioning from DNPR2 to DNPR3 is highlighted with a grey vertical line. Asterisks reflect p<0.05 for the slope of a linear model with pre- and post-transition as the independent variable. Mitigation strategies represent recoding each treatment course with the main diagnosis either unchanged, with the most “severe” diagnosis, or with the final diagnosis from the treatment course. Visits were considered part of the same treatment course if they had the same course-element-identifier (see Supplementary Table 1 for further details).

**Table 1.** Relative and absolute changes in proportions of visits belonging to each diagnostic subchapter before and after the DNPR2 to DNPR3 transition

| **Diagnostics subchapter** | **Proportion of diagnoses in DNPR2** | **Proportion of diagnoses in DNPR3** | **Absolute difference** | **Relative difference** |
| --- | --- | --- | --- | --- |
| **F0 – Organic disorders** | 1.1% | 0.1% | -1.0% | -93.7% |
| **F1 – Substance abuse** | 1.2% | 1.3% | 0.1% | 5.5% |
| **F2 – Psychotic disorders** | 15.8% | 16.8% | 0.9% | 6.0% |
| **F3 – Mood disorders** | 23.6% | 23.7% | 0.1% | 0.1% |
| **F4 – Neurotic & stress-related\*** | 17.1% | 15.1% | -2.0% | -11.8% |
| **F5 – Eating & sleeping disorders<sup>#</sup>** | 4.0% | 4.3% | 0.3% | 8.5% |
| **F6 – Personality disorders** | 8.3% | 8.4% | 0.1% | 0.7% |
| **F7 – Mental retardation** | 1.3% | 1.0% | -0.3% | -21.6% |
| **F8 – Developmental disorders** | 6.9% | 7.8% | 0.8% | 11.7% |
| **F9 – Child & adolescent disorders<sup>†</sup>** | 24.7% | 27.2% | 2.5% | 10.2% |
| *Neurotic, stress-related, and somatoform disorders<br><sup>#</sup>Behavioural syndromes associated with physiological disturbances and physical factors<br><sup>†</sup>Behavioural and emotional disorders with onset usually occurring in childhood and adolescence (F90-F98) |  |  |  |  |

Applying the mitigation strategies (most severe-or last diagnosis) resulted in destabilisation for most subchapters, creating larger breaks in the diagnostic time-series at the transition from the DNPR2 to the DNPR3 compared to the unmanipulated/unchanged data (Figure 2). Considering visits to be part of the same treatment course only if they were to the same outpatient clinic, yielded similar levels of stability (see Supplementary Figure 2). Since only outpatient visits should be affected by the change in diagnostic coding from the DNPR2 to the DNPR3, we examined the main diagnoses assigned in relation to inpatient treatment/admissions as a “negative control”. These diagnostic time-series appeared stable (see Supplementary Figure 3).

### ii) Diagnostic stability during outpatient treatment courses in the DNPR3 era

Figure 3 visualises the high diagnostic stability of the psychiatric outpatient treatment courses in the DNPR3. For more than 90% of the outpatient treatment courses, the main diagnosis assigned in relation to the first and final visit came from the same ICD-10 subchapter (see Figure 3 and Supplementary Table 2). For only three diagnostic subchapters did the first and final diagnosis match for less than 90% of the outpatient treatment courses, namely F1 - substance abuse (78.3%), F3 - mood disorders (87.6%) and F8 – developmental disorders (88.4%) (Supplementary Table 2).

**Figure 3.**
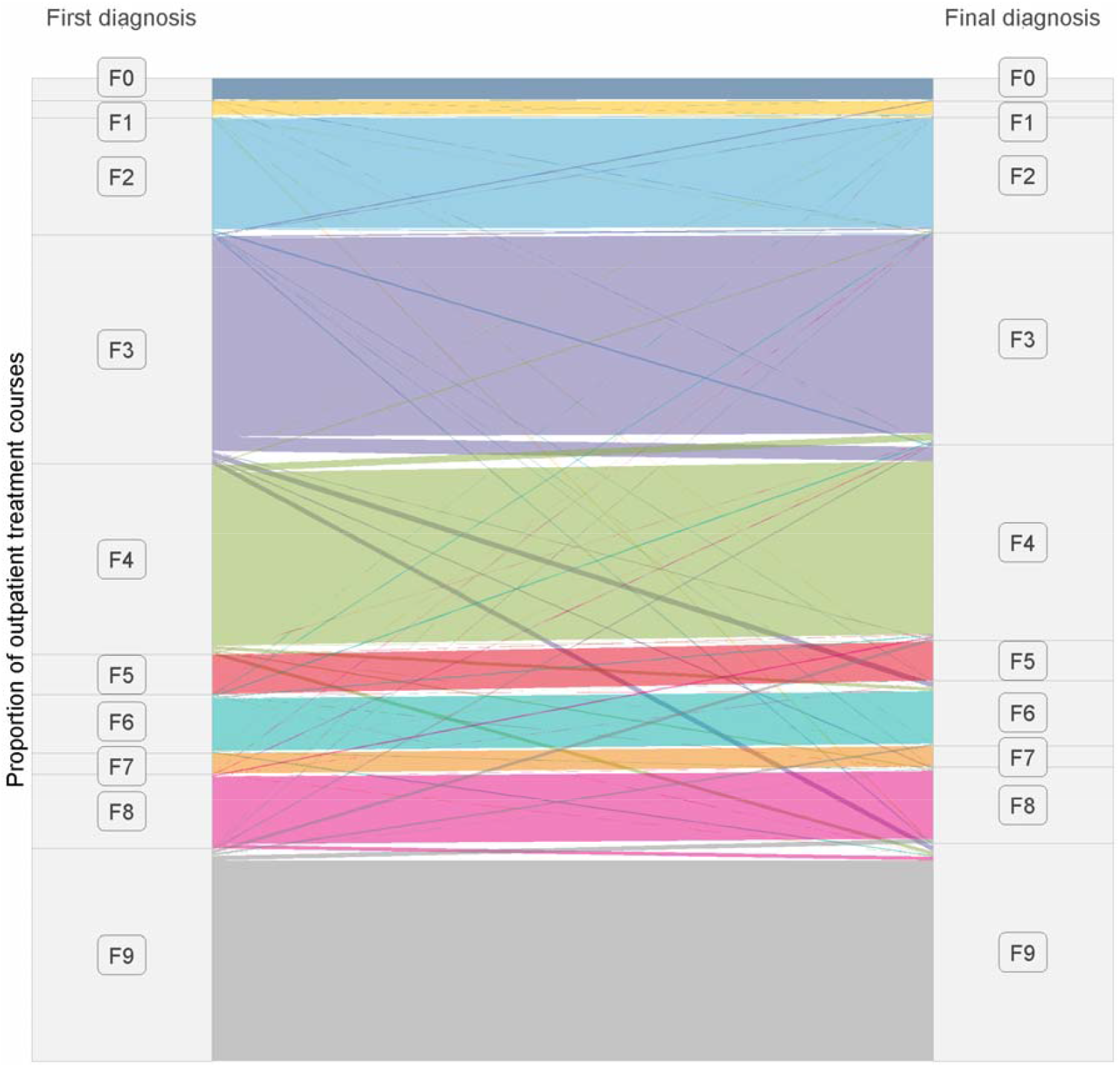
Diagnostic stability of outpatient treatment courses in the DNPR3 era Visits were considered part of the same treatment course if they had the same course-element-identifier (see Supplementary Table 1 for further detail). The thickness of lines is proportional to the number of outpatient treatment courses. Separated into first diagnosis (left) and final diagnosis (right). Colours reflect the subchapter of the final diagnosis. For exact counts and proportions, see supplementary tables 1 and 2.

### iii) Mean number of unique diagnoses per outpatient treatment course across the DNPR2-DNPR3 eras

Figure 4 shows the mean (quarterly) number of unique diagnoses per outpatient treatment course across the DNPR2-DNPR3 eras. At the FXX-level, there was no statistically significant change in the mean number of unique diagnoses per treatment course from the DNPR2 (mean = 0.75) to the DNPR3 era (mean = 0.73) (Figure 4A). However, there was larger variation between quarters during the DNPR3 era compared to the DNPR2 era (SD_Unmitigated DNPR2_ = 0.026, SD_Unmitigated DNPR3_ = 0.048). We observed periodicity, with a drop in every 3rd quarter, reflecting a lower amount of outpatient visits during the summer holiday in July/August. The lack of statistically significant changes persisted across levels of diagnostic truncation (FX, FXX.X and FXX.XX – see Figure 4A). Both mitigation strategies (most severe and final diagnosis trumping all other, respectively) worsened this contrast by excessively lowering the number of diagnoses per treatment course from the DNPR2 (mean = 0.73) to the DNPR3 era (mean_Most severe_ = 0.68, mean_Final visit_ = 0.68) (Figure 4B), resulting in statistically significant changes in means from the DNPR2 to the DNPR3. When considering visits to be part of a treatment course only when they were in the same outpatient clinic, both mitigation strategies decreased the mean number of unique diagnoses, but not to the point of statistical significance (Figure 4B).

**Figure 4.**
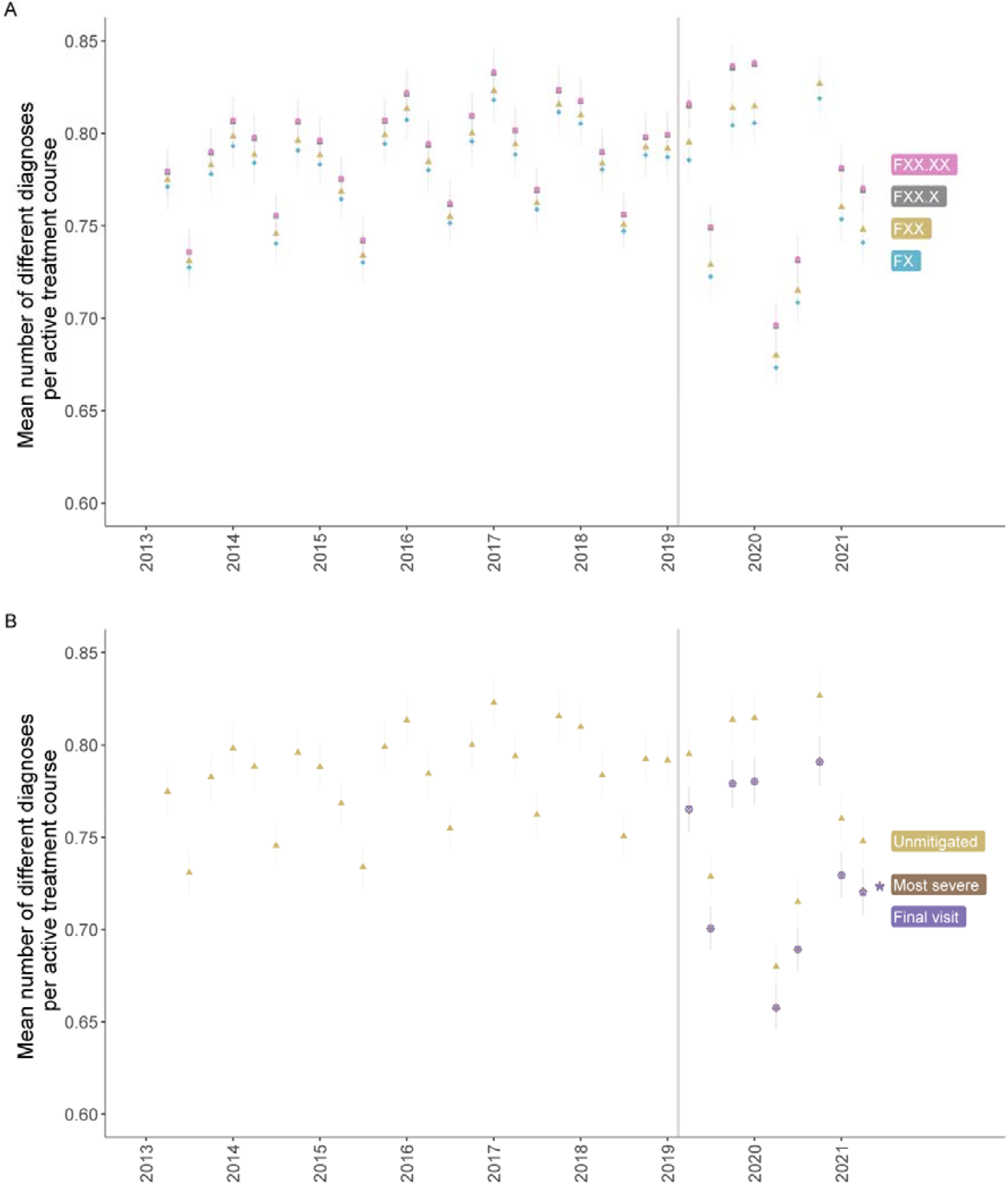
Mean number of unique psychiatric main diagnoses per active treatment course Visits were considered part of the same treatment course if they had the same DNPR3 course-element-identifier (see methods or Supplementary Table 1 for further detail). A treatment course was considered active up until 180 days after the last recorded visit. The transition date from DNPR2 to DNPR3 is marked with a grey vertical line. Asterisks reflect p < 0.05 for a t-test comparing the mans before and after the transition. Mitigation strategies represent recoding a treatment course with the main diagnosis either unchanged, with the most “severe” diagnosis, or with the *final* diagnosis from a sequence. **A:** By levels of truncation of the ICD-10 diagnostic codes and **B:** By mitigation strategy truncated at ICD-10 level FX.

## Discussion

In this cohort study based on data from all in- and outpatient visits to the Psychiatric Services of the Central Denmark Region in the period from January 1, 2013, to June 1, 2021, we showed that the transition from the DNPR2 to the DNPR3 only caused a minor destabilisation in the diagnostic time-series for the psychiatric ICD-10 subchapters. Furthermore, we found that main diagnoses assigned in relation to outpatient treatment courses in the DNPR3 era were quite stable from beginning (first visit) to end (last visit). At the level of the individual patient, there was no substantial or statistically significant change in the number of diagnoses per outpatient treatment course across the DNPR2-DNPR3 transition, but an increase in the variability in the DNPR3 era.

To our knowledge, this is the first study of the stability of diagnostic coding after the transition to the DNPR3 and it therefore contributes with novel information. The shift from the DNPR2 to the DNPR3 is the first major administrative shift of the diagnostic coding practice in Denmark since the ICD-10 replaced the ICD-8 in 1994.^1^ We have searched the literature for reports of similar administrative shifts in national patient registries, but found none. A recent review of health registries in the Nordic countries^11^ covered the validity of the diagnoses in these registries, without mentioning administrative changes, including that from the DNPR2 to the DNPR3. We therefore contacted representatives from the national patient registries in other Nordic countries (Sweden, Norway, Finland and Iceland) to inquire whether administrative shifts in diagnostic coding, comparable to that from the DNPR2-DNPR3 transition, had been made in recent times. Representatives from Norway, Finland and Iceland reported that there had been no administrative changes in diagnostic coding in the national patient registries. The representative from Sweden, however, pointed to an administrative change in 2015 regarding the registration of specialized outpatient visits. Prior to 2015, every specialized outpatient visit (e.g., to an emergency room) that subsequently led to an inpatient visit was only registered as an inpatient visit. After 2015, however, it was registered as two separate visits, one outpatient and one inpatient, leading to an increase in the number of registered outpatient visits. Thus, if basing an analysis solely on data from outpatient visits in Sweden, the 2015 administrative change would lead to an increase in the number of “cases” without an actual increase in morbidity.^12^

Overall, our analyses showed that the change in diagnostic coding accompanying the transition from the DNPR2 to the DNPR3 only had a minor impact on the stability of the diagnostic time-series as well as on the population-adjusted within-quarter incidence of psychiatric diagnoses. This should offer some optimism regarding the validity of future psychiatric epidemiological studies using the DNPR as data source. We did, however, observe increased variability in the number of unique psychiatric main diagnosis per active treatment course after the transition from the DNPR2 to the DNPR3. For an increase in variance to occur without a change in the mean, there must be a simultaneous increase i) in the proportion of treatment courses with a high number of diagnoses, and ii) in the proportion of treatment courses with a low number of diagnoses. In fact, the COVID-19 pandemic, which coincided with the observed increased variability, may have led to such a change in the pattern of contacts/diagnoses. Studies from Danish Psychiatric Services have suggested that the COVID-19 pandemic has caused some patients to exhibit new symptoms (e.g., anxiety),^13–15^ while also resulting in patients withdrawing from (or their appointments being cancelled by) the healthcare system, leading to fewer diagnoses.^16,17^ If this is the explanation, the increased variability may be transitory and return to the base level as the pandemic comes to an end. Other societal- and patient-level changes than the COVID-19 pandemic, leading to the same contact/diagnostic pattern, could, however, also have played a role regarding the observed variability.

There are limitations to this study, which must be considered. First, we only had access to data from electronic patient records in the Central Denmark Region, provided by the regional Business Intelligence Office. While this is ideal for the purpose of ensuring diagnostic stability in relation to research based on this data source,^9^ one could argue that it is not identical to data from the DNPR. However, given that the DNPR and the Business Intelligence Office in the Central Denmark Region receive data from the same source, this is unlikely to affect our results. Second, and relatedly, the data for the present study stems solely from the Central Denmark Region, which could make the results less generalisable.

Nevertheless, since reporting of diagnoses is based on national standards, it is likely to generalise across the Danish regions. Third, this study only focused on the diagnostic stability in relation to treatment in the Psychiatric Services. However, since the change in diagnostic coding practice accompanying the transition from the DNPR2 to the DNPR3 was the same across all medical and surgical specialities in the Danish secondary healthcare sector, our results are likely to generalise to those as well, but this should be subjected to empirical testing. Fourth, while the main analytical level in this study, namely the F-subchapters (FX), captures much information, there is heterogeneity within these chapters, one example being F32/33 - unipolar depression and F30/F31 - mania/bipolar disorder. It was, however, beyond the scope of this study to investigate the stability of each of the 71 different FXX diagnoses in the ICD-10. Research groups working with data from the DNPR having particular emphasis on specific diagnoses should probably investigate the stability of those across the DNPR2-DNPR3 transition, as our results do not necessarily generalise to all FXX diagnoses (nor to diagnoses at even greater granular level, e.g., FXX.X and FXX.XX).

In conclusion, the change in diagnostic coding of psychiatric outpatient visits accompanying the transition from the DNPR2 to the DNPR3 did not appear to lead to substantial breaks in the diagnostic time-series represented by the DNPR – at least not at the diagnostic subchapter level. Therefore, the change from the DNPR2 to the DNPR3 is unlikely to pose a substantial threat to the validity of most psychiatric research based on this data source.

## Data Availability

Due to restrictions in Danish law for protecting patient privacy, the data used in this study is only available for research projects conducted by employees in the Central Denmark Region following approval from the Legal Office under the Central Denmark Region (in accordance with the Danish Health Care Act 46, Section 2). However, similar data can be accessed through Statistics Denmark. Danish institutions can apply for authorization to work with data within Statistics Denmark, and such organisations can provide access to affiliated researchers inside and outside of Denmark.

## Acknowledgements

The authors thank Bettina Nørremark from Aarhus University Hospital – Psychiatry for her assistance with extraction of data, and Elisabeth Flebbe from Aarhus University Hospital – Psychiatry for discussions of diagnostic coding practice across the DNPR2 and DNPR3.

## Funding

The study is supported by grants from the Lundbeck Foundation (grant number: R344-2020-1073), the Danish Cancer Society (grant number: R283-A16461), the Central Denmark Region Fund for Strengthening of Health Science (grant number: 1-36-72-4-20) and the Danish Agency for Digitisation Investment Fund for New Technologies (grant number 2020-6720) to Østergaard, who reports further funding from the Lundbeck Foundation (grant number: R358-2020-2341), the Novo Nordisk Foundation (grant number: NNF20SA0062874) and Independent Research Fund Denmark (grant number: 7016-00048B). The funders played no role in study design, collection, analysis or interpretation of data, the writing of the report or the decision to submit the paper for publication.

## Conflicts of interest

Danielsen has received a speaker honorarium from Otsuka Pharmaceutical. Østergaard received the 2020 Lundbeck Foundation Young Investigator Prize. Furthermore, Østergaard owns units of mutual funds with stock tickers DKIGI and WEKAFKI, as well as units of exchange-traded funds with stock ticker TRET and EUNL.

## Notes

### Competing Interest Statement

Danielsen has received a speaker honorarium from Otsuka Pharmaceutical. Oestergaard received the 2020 Lundbeck Foundation Young Investigator Prize. Furthermore, Oestergaard owns units of mutual funds with stock tickers DKIGI and WEKAFKI, as well as units of exchange-traded funds with stock tickers QDVH, EUNL and SADM.

### Author Declarations

Central Denmark Region Legal Office of the Central Denmark Region waived ethical approval for this work (1-10-72-1-22).

