## Supplementary for "Stability of diagnostic coding of psychiatric outpatient visits across the transition from the second to the third version of the Danish National Patient Registry"

###### Supplementary Table 1. Differences between a DNPR2 course-identifier and a DNPR3 course-element-identifier

|  | **DNPR2** | **DNPR3** |
| --- | --- | --- |
| **Allowed visit types in one course** | (Outpatient visits and phone/video consultations) OR (inpatient visits). | All types: phone/video-consultations, outpatient- and inpatient visits. |
| **Allowed geographical location in one course** | The same subsection^+^ | The same department^#^ |
| **Other triggers for creating a new course** | Significant change in main diagnosis^&^ OR transition from fast-track diagnostic course to regular outpatient course |  |
| ^+^E.g. Clinic for Anxiety and OCD, Clinic for Depression and Mania or The Psychiatric Diagnostic Unit.  ^#^E.g. Department for Depression and Anxiety, Department for Psychoses or Department of Forensic Psychiatry.  **^&^A change at the level of FX, e.g. from F2 to F4.** | | |

###### Supplementary Figure 1. Proportion of outpatients in a quarter with a within-quarter incident main diagnosis by ICD-10 subchapter

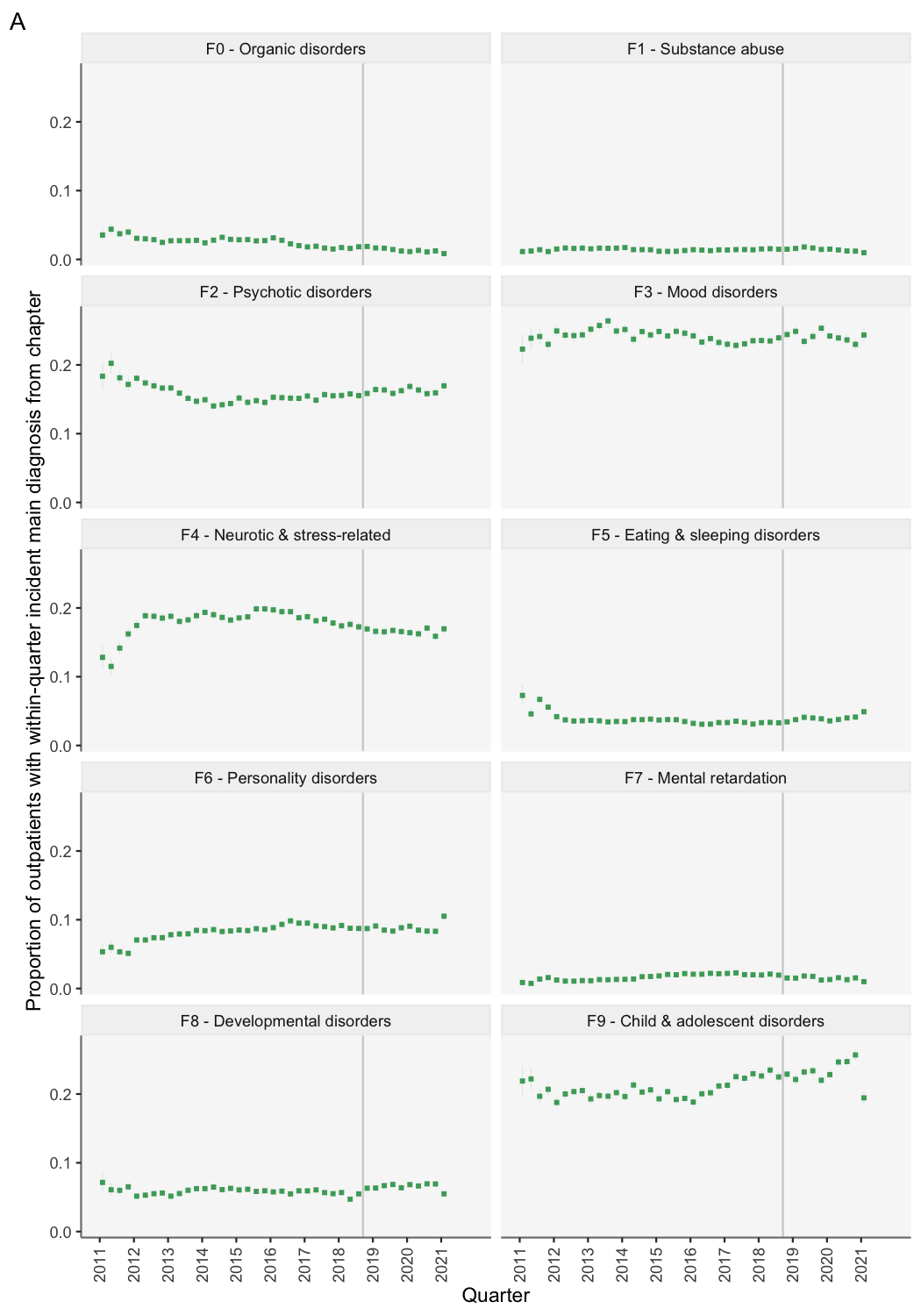

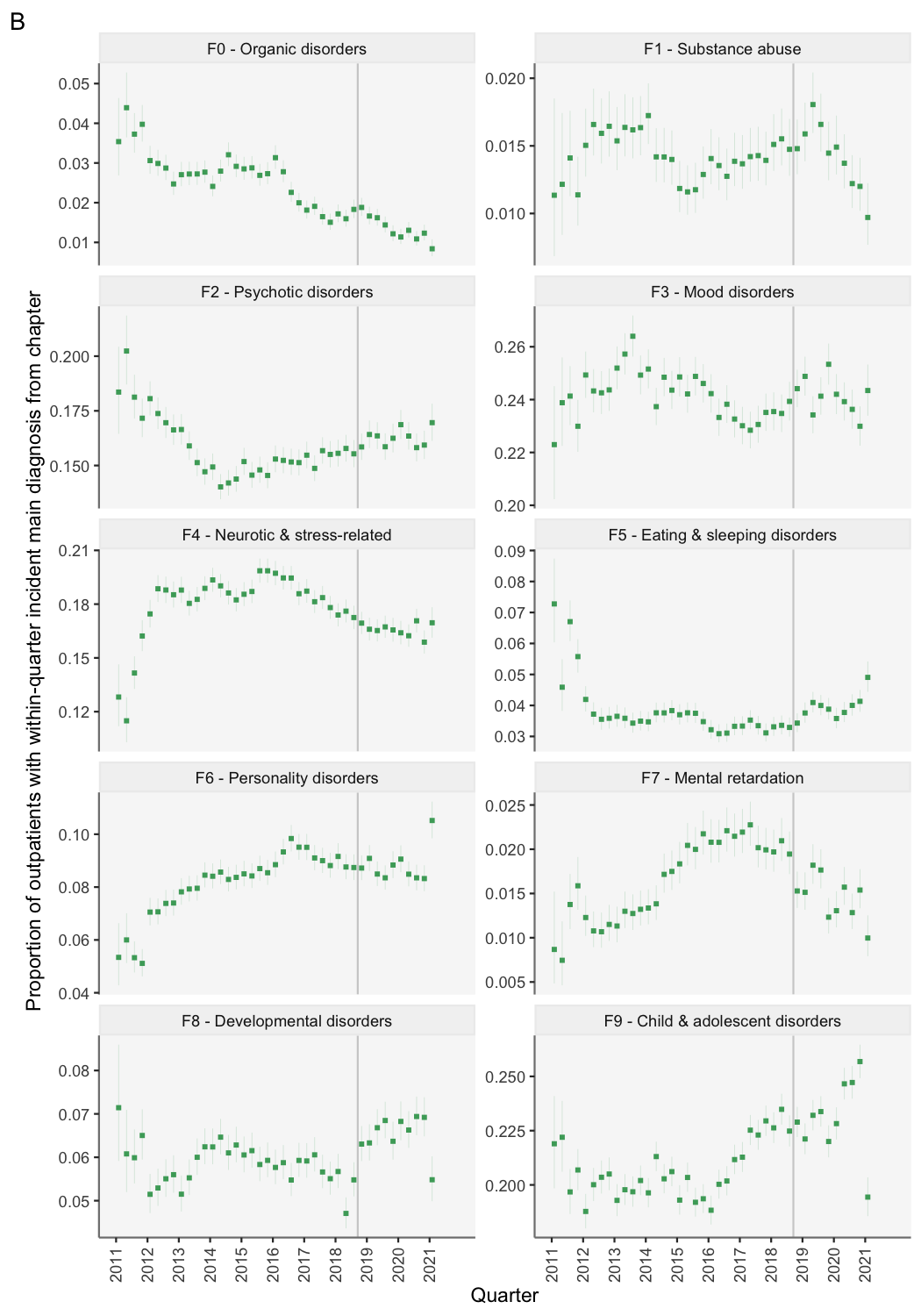

Line-ranges represent 95% confidence intervals. The date of transitioning from DNPR2 to DNPR3 is highlighted with a grey line. **A:** y-scale is standardised. **B:** y-scale is allowed to vary between panels.

**Supplementary Figure 2.** Proportion of outpatients in each quarter with a within-quarter incident main diagnosis from the respective ICD-10 subchapters. Visits were considered to be part of the same treatment course only if they were to the same outpatient clinic.

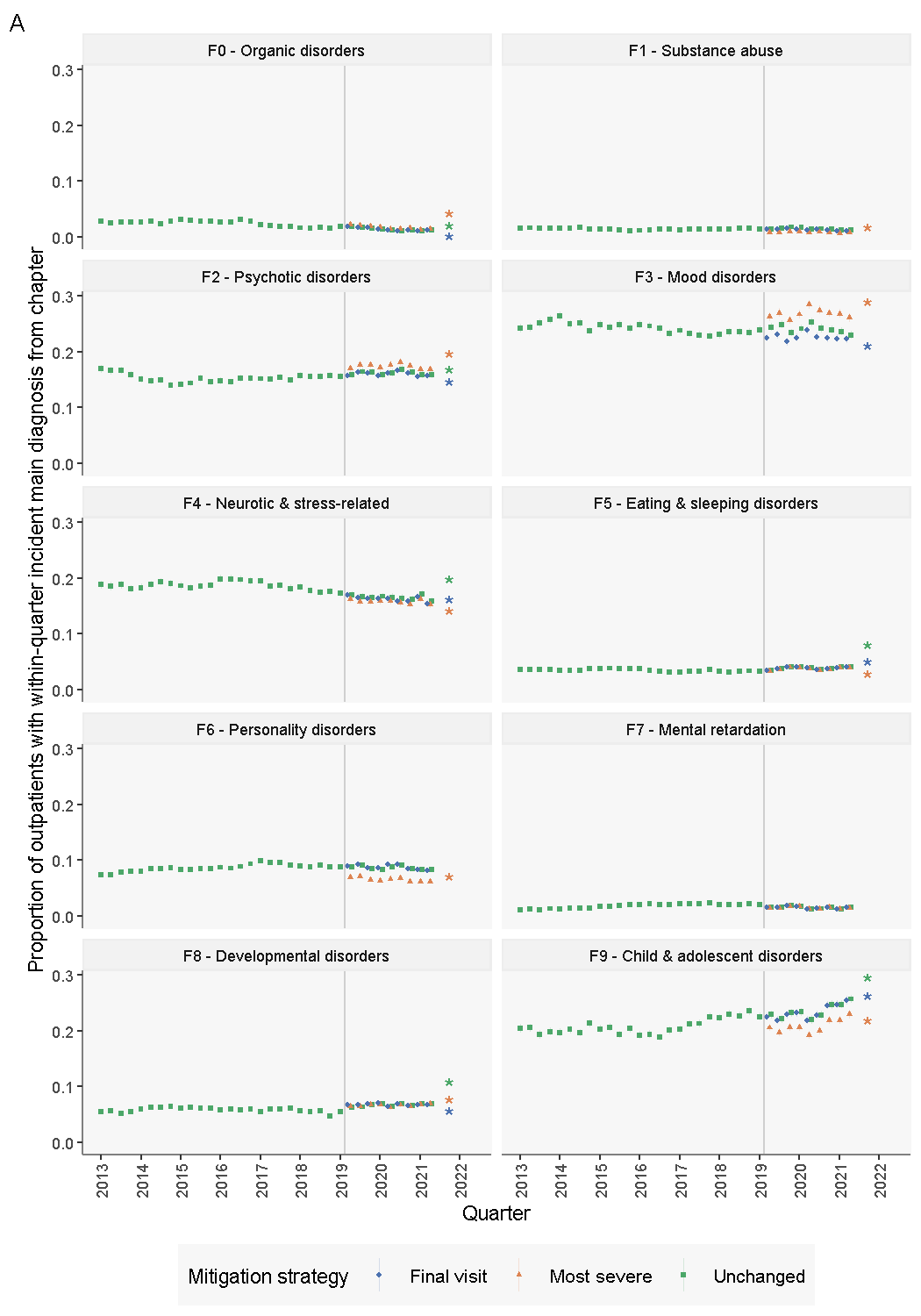

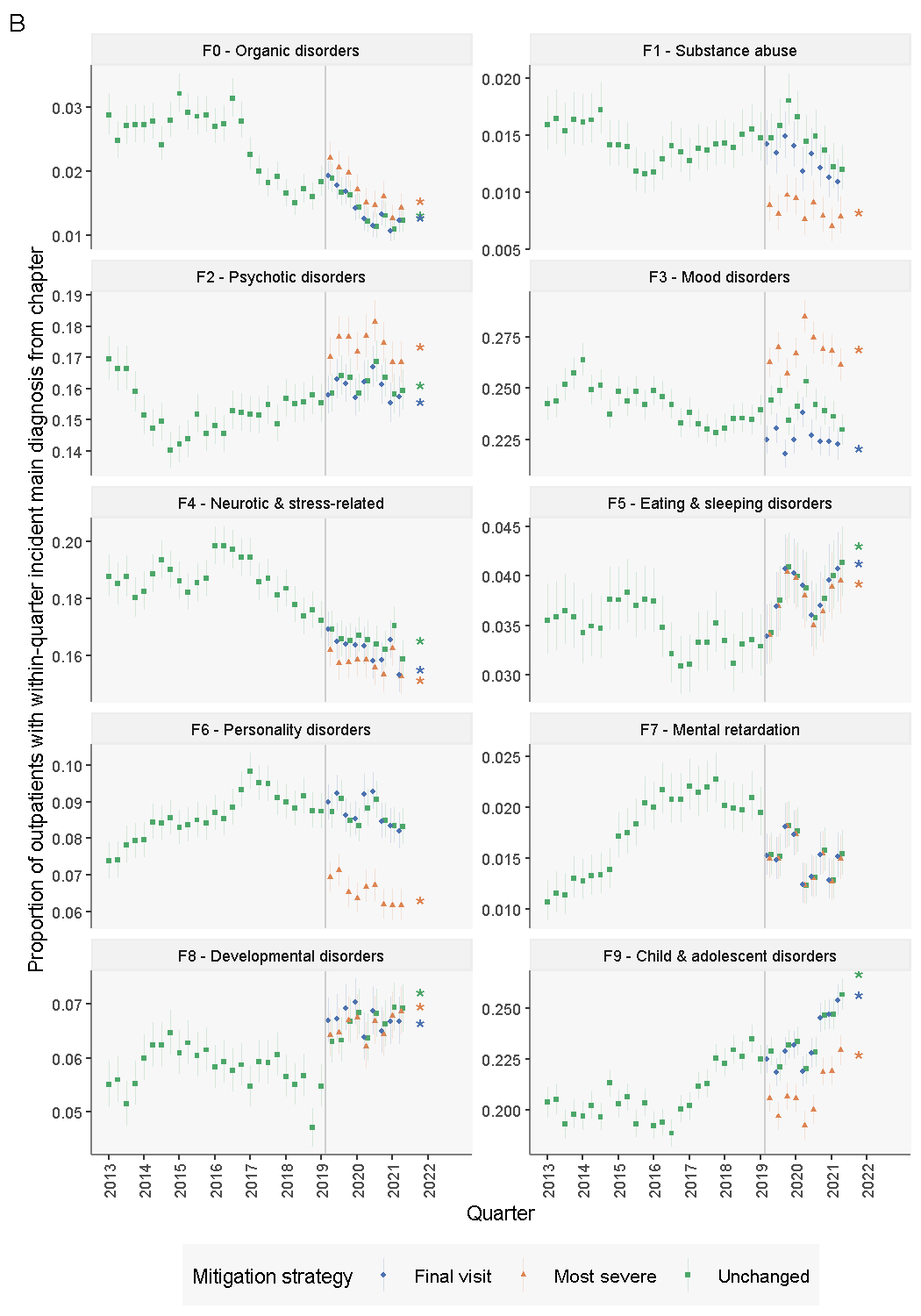

Line-ranges represent 95% confidence intervals. The date of transitioning from the DNPR2 to the DNPR3 is highlighted with a grey line. Mitigation strategies represent recoding each treatment course with the main diagnosis either unchanged, with the most “severe” diagnosis, or with the final diagnosis from the treatment course. Asterisks reflect p < 0.05 for a difference in the mean proportion comparing before and after the DNPR-transition. **A:** y-scale is standardised. **B:** y-scale is allowed to vary between panels.

##### Supplementary Figure 3. Proportion of inpatients in a quarter that received a main diagnosis from the respective ICD-10 subchapters

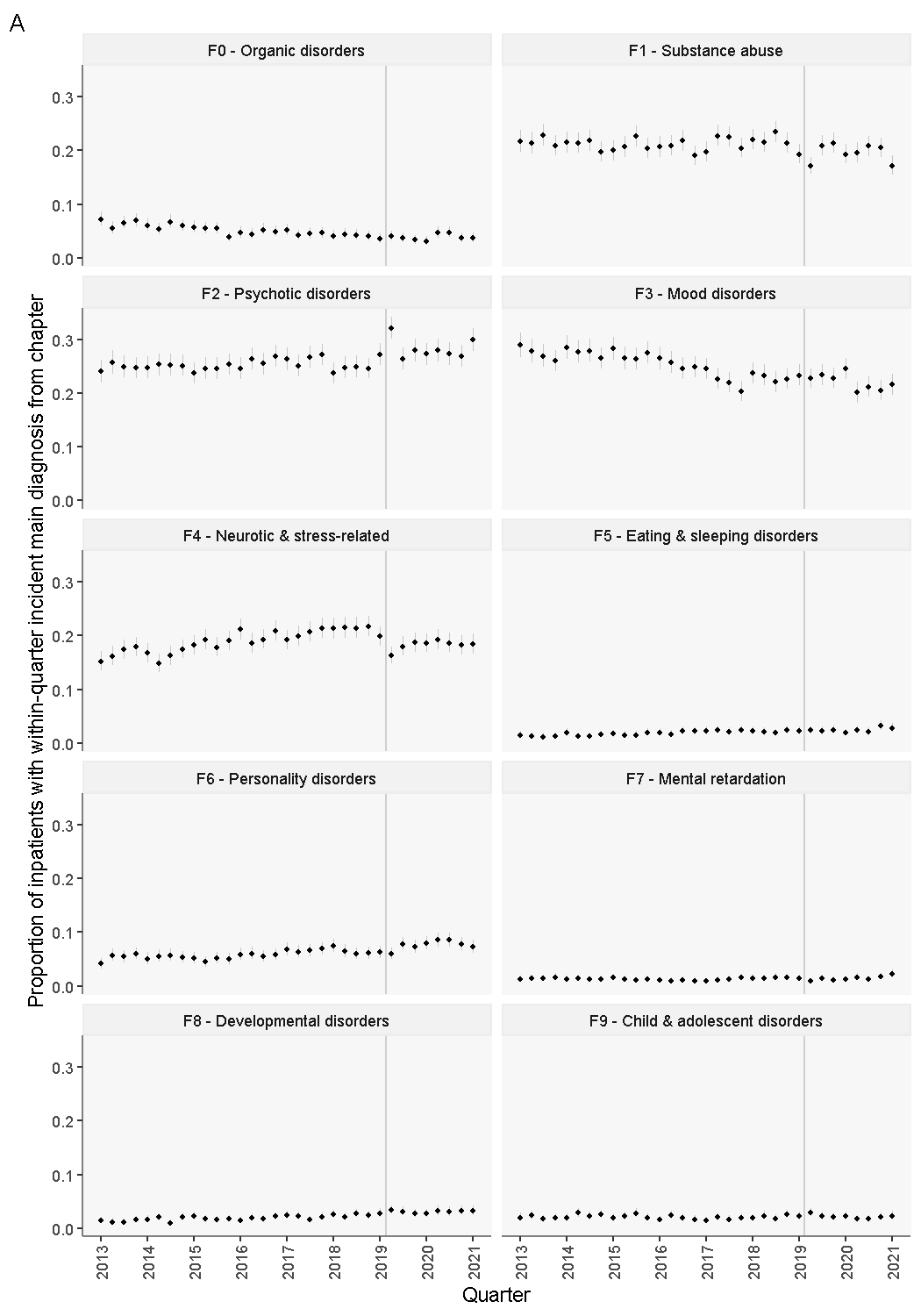

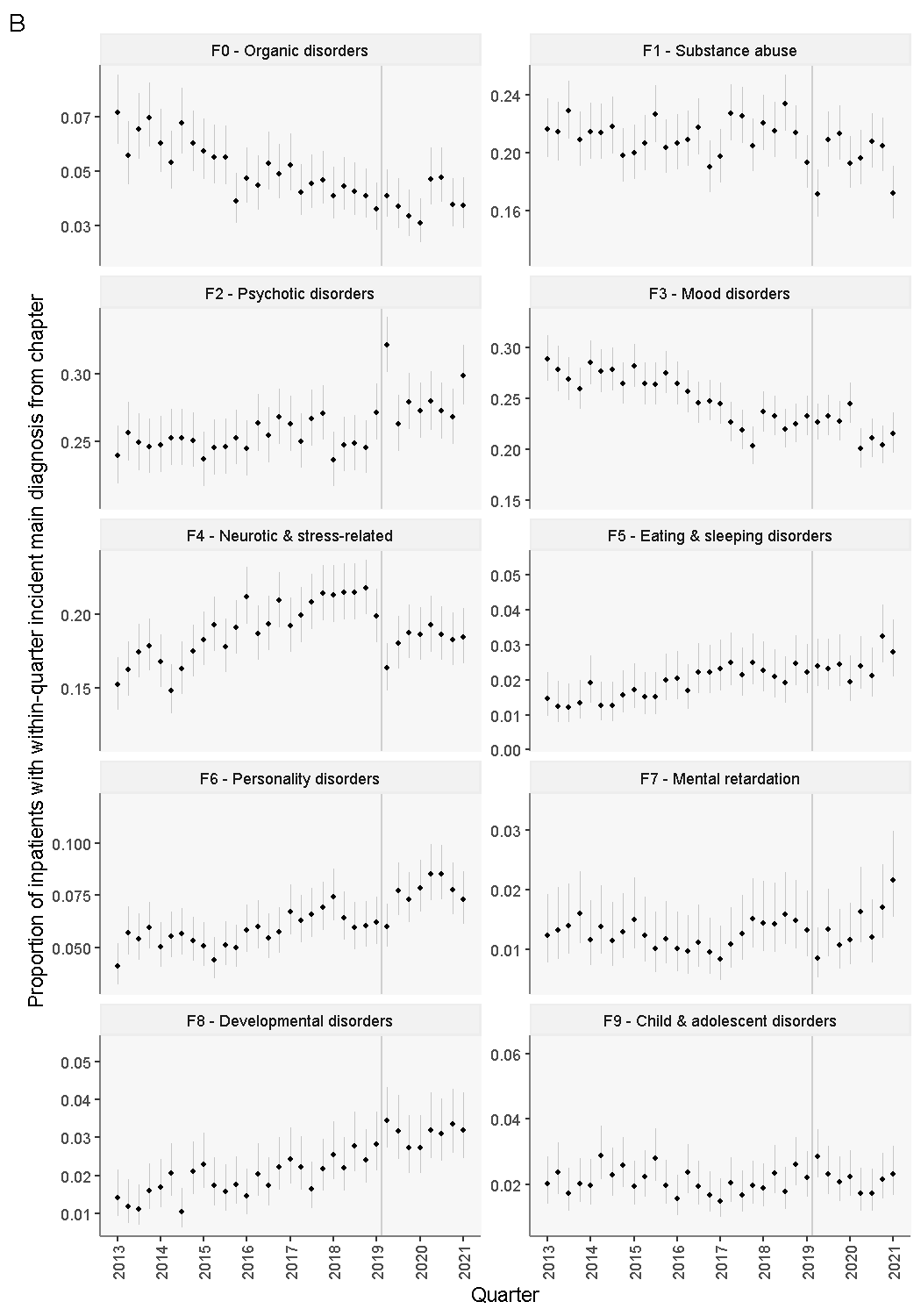

Line-ranges represent 95% confidence intervals. The date of transitioning from DNPR2 to DNPR3 is highlighted with a grey line. Mitigation strategies represent recoding a treatment course with the main diagnosis either unchanged, from the final visit, or from the lowest-digit ICD-10 subchapter. A: y-scale is standardised. B: y-scale is allowed to vary between panels.

###### Supplementary Figure 4. Mean number of unique psychiatric main diagnoses per active treatment course

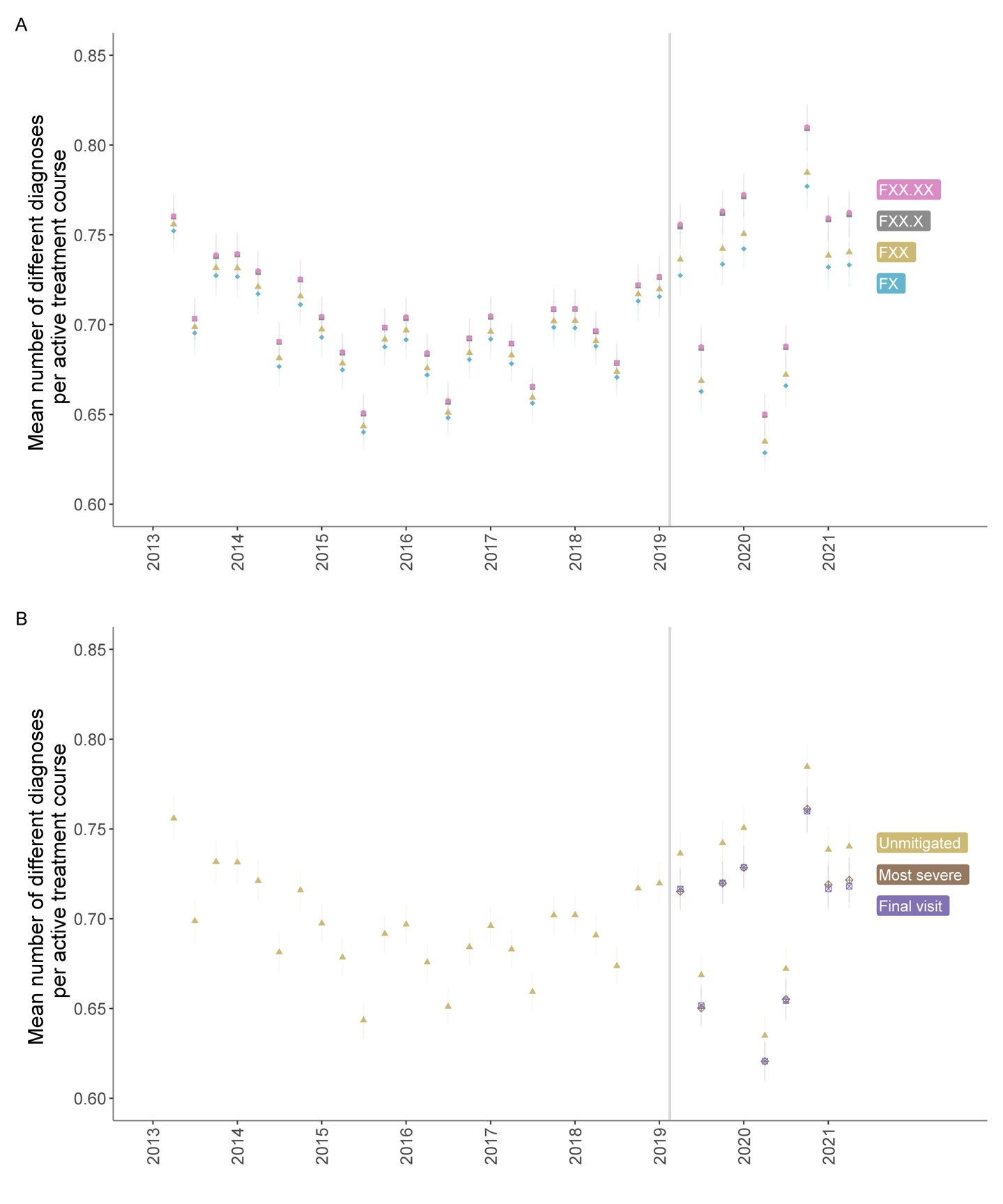

Visits are considered a treatment course only if they are both i) from the same patient and b) in the same outpatient clinic. A treatment course terminates 180 days after the latest visit. The transition date from DNPR2 to DNPR3 is marked with a grey vertical line. Asterisks reflect p < 0.05 for a t-test of the means before and after the transition. Mitigation strategies represent recoding a treatment course with the main diagnosis either unchanged, with the most “severe” diagnosis, or with the diagnosis from the *final* visit. **A:** By levels of truncation of the ICD-10 diagnostic codes and **B:** By mitigation strategy truncated to ICD-10 level FXX.

**Supplementary Table 2.** Treatment courses grouped by first diagnosis

| First Diagnosis | Final Diagnosis | n | % of First Diagnosis |
| --- | --- | --- | --- |
| F0 | **F0** | **807** | **95.1%** |
|  | F1 | 6 | 0.7% |
|  | F2 | 9 | 1.1% |
|  | F3 | 16 | 1.9% |
|  | F4 | 6 | 0.7% |
|  | F7 | 5 | 0.6% |
| F1 | **F1** | **520** | **80.1%** |
|  | F2 | 30 | 4.6% |
|  | F3 | 22 | 3.4% |
|  | F4 | 10 | 1.5% |
|  | F6 | 19 | 2.9% |
|  | F9 | 41 | 6.3% |
|  | Other | 7 | 1.1% |
| F2 | F0 | 8 | 0.2% |
|  | F1 | 31 | 0.7% |
|  | **F2** | **4256** | **94.1%** |
|  | F3 | 40 | 0.9% |
|  | F4 | 71 | 1.6% |
|  | F6 | 25 | 0.6% |
|  | F7 | 9 | 0.2% |
|  | F8 | 29 | 0.6% |
|  | F9 | 53 | 1.2% |
| F3 | F0 | 44 | 0.5% |
|  | F1 | 24 | 0.3% |
|  | F2 | 56 | 0.7% |
|  | **F3** | **7423** | **87.8%** |
|  | F4 | 488 | 5.8% |
|  | F5 | 22 | 0.3% |
|  | F6 | 189 | 2.2% |
|  | F8 | 48 | 0.6% |
|  | F9 | 160 | 1.9% |
| F4 | F0 | 9 | 0.1% |
|  | F1 | 13 | 0.2% |
|  | F2 | 38 | 0.6% |
|  | F3 | 247 | 3.7% |
|  | **F4** | **6091** | **91.1%** |
|  | F5 | 10 | 0.1% |
|  | F6 | 118 | 1.8% |
|  | F8 | 50 | 0.7% |
|  | F9 | 112 | 1.7% |
| F5 | F3 | 10 | 0.7% |
|  | F4 | 12 | 0.9% |
|  | **F5** | **1350** | **96.7%** |
|  | F6 | 8 | 0.6% |
|  | F8 | 10 | 0.7% |
|  | F9 | 6 | 0.4% |
| F6 | F1 | 12 | 0.5% |
|  | F2 | 36 | 1.6% |
|  | F3 | 43 | 2% |
|  | F4 | 34 | 1.5% |
|  | **F6** | **2018** | **91.6%** |
|  | F8 | 16 | 0.7% |
|  | F9 | 38 | 1.7% |
|  | Other | 5 | 0.2% |
| F7 | F2 | 9 | 1.2% |
|  | F3 | 5 | 0.6% |
|  | F4 | 5 | 0.6% |
|  | **F7** | **742** | **95.1%** |
|  | F9 | 14 | 1.8% |
|  | Other | 5 | 0.6% |
| F8 | F2 | 11 | 0.4% |
|  | F3 | 17 | 0.6% |
|  | F4 | 49 | 1.9% |
|  | F5 | 6 | 0.2% |
|  | F7 | 7 | 0.3% |
|  | **F8** | **2429** | **92.3%** |
|  | F9 | 108 | 4.1% |
|  | Other | 5 | 0.2% |
| F9 | F1 | 12 | 0.2% |
|  | F2 | 27 | 0.3% |
|  | F3 | 37 | 0.5% |
|  | F4 | 94 | 1.2% |
|  | F6 | 76 | 1% |
|  | F7 | 8 | 0.1% |
|  | F8 | 162 | 2% |
|  | **F9** | **7511** | **94.8%** |
| To comply with Danish Data Legislation, rows with n < 5 were collapsed into “other”. If the “other” category had n < 5, it was removed prior to calculation of percentages. | | | |

**Supplementary Table 3.** Treatment courses grouped by final diagnosis

| First Diagnosis | Final Diagnosis | n | % of Last Diagnosis |
| --- | --- | --- | --- |
| F0 | **F0** | **807** | **93%** |
| F2 |  | 8 | 0.9% |
| F3 |  | 44 | 5.1% |
| F4 |  | 9 | 1% |
| F0 | **F1** | 6 | 1% |
| F1 |  | **520** | **84.1%** |
| F2 |  | 31 | 5% |
| F3 |  | 24 | 3.9% |
| F4 |  | 13 | 2.1% |
| F6 |  | 12 | 1.9% |
| F9 |  | 12 | 1.9% |
| F0 | **F2** | 9 | 0.2% |
| F1 |  | 30 | 0.7% |
| F2 |  | **4256** | **95.2%** |
| F3 |  | 56 | 1.3% |
| F4 |  | 38 | 0.8% |
| F6 |  | 36 | 0.8% |
| F7 |  | 9 | 0.2% |
| F8 |  | 11 | 0.2% |
| F9 |  | 27 | 0.6% |
| F0 | **F3** | 16 | 0.2% |
| F1 |  | 22 | 0.3% |
| F2 |  | 40 | 0.5% |
| F3 |  | **7423** | **94.4%** |
| F4 |  | 247 | 3.1% |
| F5 |  | 10 | 0.1% |
| F6 |  | 43 | 0.5% |
| F7 |  | 5 | 0.1% |
| F8 |  | 17 | 0.2% |
| F9 |  | 37 | 0.5% |
| F0 | **F4** | 6 | 0.1% |
| F1 |  | 10 | 0.1% |
| F2 |  | 71 | 1% |
| F3 |  | 488 | 7.1% |
| F4 |  | **6091** | **88.8%** |
| F5 |  | 12 | 0.2% |
| F6 |  | 34 | 0.5% |
| F7 |  | 5 | 0.1% |
| F8 |  | 49 | 0.7% |
| F9 |  | 94 | 1.4% |
| F3 | **F5** | 22 | 1.6% |
| F4 |  | 10 | 0.7% |
| F5 |  | **1350** | **97.3%** |
| F8 |  | 6 | 0.4% |
| F1 | **F6** | 19 | 0.8% |
| F2 |  | 25 | 1% |
| F3 |  | 189 | 7.7% |
| F4 |  | 118 | 4.8% |
| F5 |  | 8 | 0.3% |
| F6 |  | **2018** | **82.3%** |
| F9 |  | 76 | 3.1% |
| F0 | **F7** | 5 | 0.6% |
| F2 |  | **9** | **1.2%** |
| F7 |  | 742 | 96.2% |
| F8 |  | 7 | 0.9% |
| F9 |  | 8 | 1% |
| F2 | **F8** | 29 | 1.1% |
| F3 |  | 48 | 1.7% |
| F4 |  | 50 | 1.8% |
| F5 |  | 10 | 0.4% |
| F6 |  | **16** | **0.6%** |
| F8 |  | 2429 | 88.5% |
| F9 |  | 162 | 5.9% |
| F1 | **F9** | 41 | 0.5% |
| F2 |  | 53 | 0.7% |
| F3 |  | 160 | 2% |
| F4 |  | 112 | 1.4% |
| F5 |  | 6 | 0.1% |
| F6 |  | 38 | 0.5% |
| F7 |  | 14 | 0.2% |
| F8 |  | **108** | **1.3%** |
| F9 |  | 7511 | 93.4% |
| F1 | **Other** | 7 | 31.8% |
| F6 |  | 5 | 22.7% |
| F7 |  | 5 | 22.7% |
| F8 |  | 5 | 22.7% |
| To comply with Danish Data Legislation, rows with n < 5 were collapsed into “other”. If the “other” category had n < 5, it was removed prior to calculation of percentages. | | | |
